# Circulating tumor DNA in neoadjuvant treated breast cancer reflects response and survival

**DOI:** 10.1101/2020.02.03.20019760

**Authors:** Mark Jesus M. Magbanua, Lamorna Brown-Swigart, Hsin-Ta Wu, Gillian L. Hirst, Christina Yau, Denise M. Wolf, Antony Tin, Raheleh Salari, Svetlana Shchegrova, Hemant Pawar, Amy L. Delson, Angela DeMichele, Minetta C. Liu, A. Jo Chien, Smita Asare, Cheng-Ho J. Lin, Paul Billings, Alexey Aleshin, Himanshu Sethi, Maggie Louie, Bernhard Zimmermann, Laura J. Esserman, Laura J. van ’t Veer

## Abstract

Pathologic complete response (pCR) to neoadjuvant chemotherapy (NAC) is strongly associated with favorable outcome. We examined the utility of serial circulating tumor DNA (ctDNA) testing for predicting pCR and risk of metastatic recurrence in 84 high-risk early breast cancer patients treated in the neoadjuvant I-SPY 2 TRIAL. Cell-free DNA (cfDNA) was isolated from 291 plasma samples collected at pretreatment (T0), 3 weeks after initiation of paclitaxel (T1), between paclitaxel and anthracycline regimens (T2), or prior to surgery (T3). A personalized ctDNA test was designed to detect 16 patient-specific mutations (from whole exome sequencing of pretreatment tumor) in cfDNA by ultra-deep sequencing. At T0, 61 of 84 (73%) patients were ctDNA-positive, which decreased over time (T1-35%; T2-14%; T3-9%). Patients who remained ctDNA-positive at T1 were significantly more likely to have residual disease after NAC (83% non-pCR) compared to those who cleared ctDNA (52% non-pCR; OR 4.33, P=0.012). After NAC, all patients who achieved pCR were ctDNA-negative (n=17, 100%). For those who did not achieve pCR (n=43), ctDNA-positive patients (14%) had significantly increased risk of metastatic recurrence (HR 10.4; 95% CI, 2.3–46.6); interestingly, patients who did not achieve pCR but were ctDNA-negative (86%) had excellent outcome, similar to those who achieved pCR (HR 1.4; 95% CI, 0.15–13.5). Lack of ctDNA clearance was a significant predictor of poor response and metastatic recurrence, while clearance was associated with improved survival regardless of pCR status. Personalized monitoring of ctDNA during NAC may aid in real-time assessment of treatment response and help fine-tune pCR as a surrogate endpoint of survival.

## Introduction

Circulating tumor ctDNA (ctDNA) in blood offers a minimally invasive approach for disease monitoring and evaluation of response to therapy *[1-3]*. Findings from recent clinical studies have shown that ctDNA may play a role in detecting minimal residual disease (MRD) and emerging therapy resistance, i.e., molecular relapse (MR), in early stage breast cancers *[4-7]*, as well as in monitoring of disease progression in patients with advanced breast cancer *[8-10]*. However, it is not yet known if failure to clear ctDNA during therapy could provide guidance for escalation of treatment to prevent early disease recurrence *[11]*.

Neoadjuvant chemotherapy (NAC) has become a standard-of-care for patients with locally-advanced breast cancer *[12]*. First, NAC provides a unique opportunity for real-time monitoring of tumor response and evaluation of drug efficacy *[13-15]*. Second, NAC may downstage tumors and thus improve chances of breast-conserving surgery *[12, 16, 17]*. Third, response to NAC provides prognostic information which can supplement those derived from standard clinico-pathologic characteristics of the primary tumor, such as subtype, nodal status, and grade *[12, 16-20]*.

Pooled analysis by Cortazar and colleagues has shown that patients who achieved a pathologic complete response (pCR, or the absence of residual cancer in the breast and lymph nodes after NAC) have significant survival advantage over those who did not *[21]*. Standard NAC have resulted in pCR for 10-50% of patients depending on subtype *[21]*. Data from the I-SPY 2 TRIAL, a multicenter phase 2 trial that evaluates investigational drugs in combination with standard NAC (paclitaxel followed by anthracycline treatment) *[22]*, have shown that pCR in women with molecularly high-risk stage II or III tumors, whether from standard or targeted therapies, unequivocally conferred a survival advantage (hazard ratio of 0.2) [23].

While pCR accurately identifies patients with low risk of relapse, studies have shown that predicting early metastatic recurrence in those with residual disease (non-pCR) is less robust *[21, 23]*. For example, survival analysis in the I-SPY 2 TRIAL (median follow-up of 3.8 years) showed that the 3-year distant disease-free survival (DRFS) of patients who achieved pCR was 95% [23]. In contrast, only 22% of non-pCR patients experienced metastatic recurrence. In this study, we evaluated the potential role of ctDNA as a biomarker for monitoring of response to NAC and assessed the additive value of ctDNA to further stratify patients with residual disease to predict early metastatic recurrence. We performed a correlative study in the I-SPY 2 TRIAL to detect ctDNA in serial plasma samples collected before, during and after NAC [24]. We used a previously analytically validated personalized ctDNA test composed of a panel of 16 most clonal single nucleotide variants (SNVs) present in the pretreatment tumor *[10, 25-27]*. The test involves multiplex polymerase chain reaction amplification followed by ultra-deep sequencing to detect tumor-specific mutations (i.e., ctDNA) in cell-free DNA (cfDNA). This approach enables more accurate monitoring of disease burden than pre-fixed driver mutation panels, as each test reflects tumor heterogeneity at the individual patient level *[5, 8, 28]*. The test is also highly sensitive, with a limit of detection of <0.01% variant allele frequency (VAF), equivalent to one mutant molecule in a background of 10,000 wildtype copies *[10, 25-27]*.

## Results

### ctDNA analysis in I-SPY 2 TRIAL patients

We performed ctDNA analysis on samples collected from 151 I-SPY 2 TRIAL patients who received standard NAC alone (n=57) or combined with MK-2206 (AKT inhibitor) treatment (n=94) (**Fig 1A**). Of the 151 patients, 61 did not have sufficient tissue or plasma samples and were excluded from the study (**Fig 1B**). Primary tumor samples from the remaining 90 patients were subjected to whole exome sequencing (WES) (**Fig 1C, Supplementary Fig S1**); and of these, 6 were excluded due to poor quality sequencing data, resulting in an analytic cohort of 84 patients.

**Fig 1.**
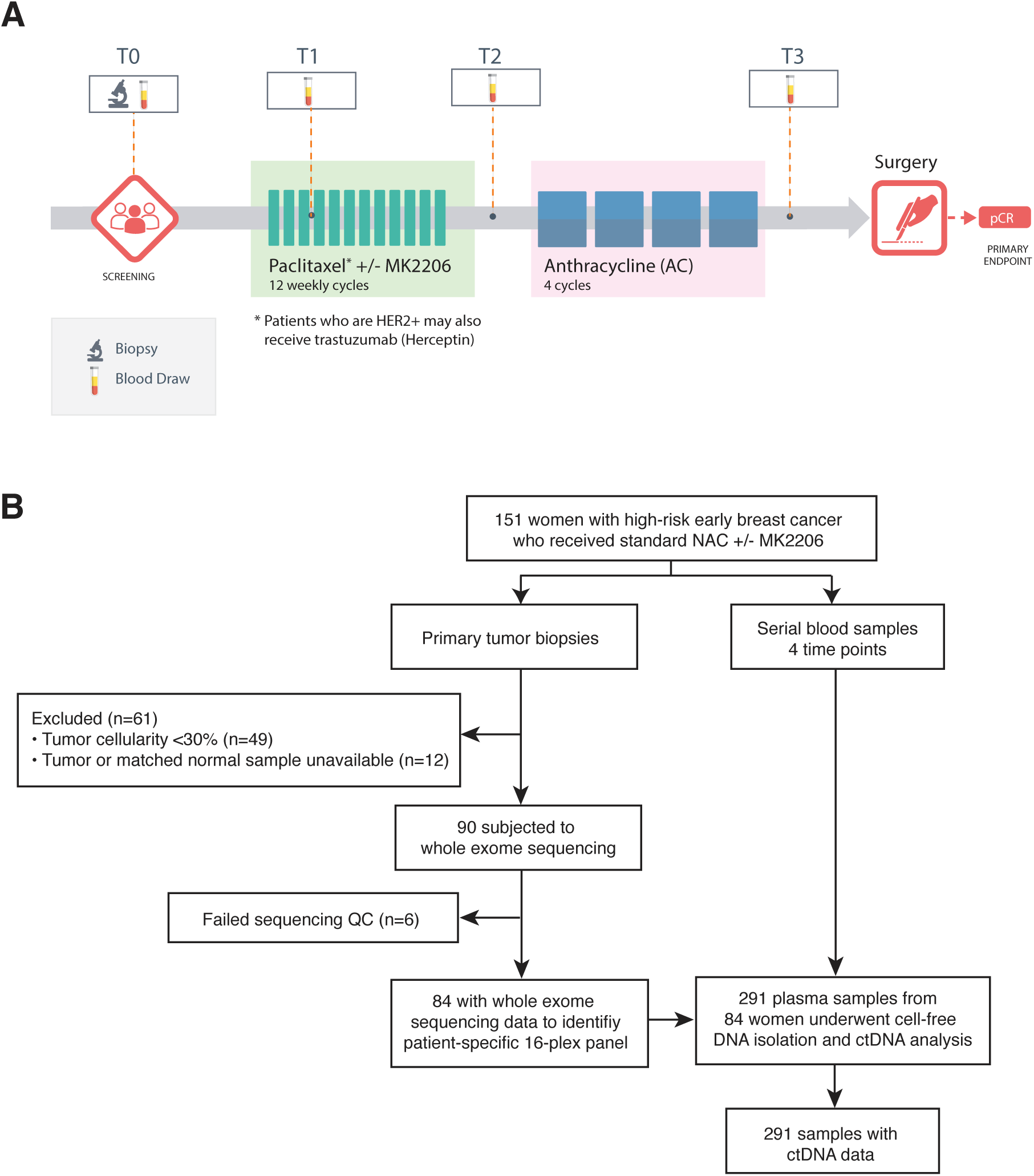
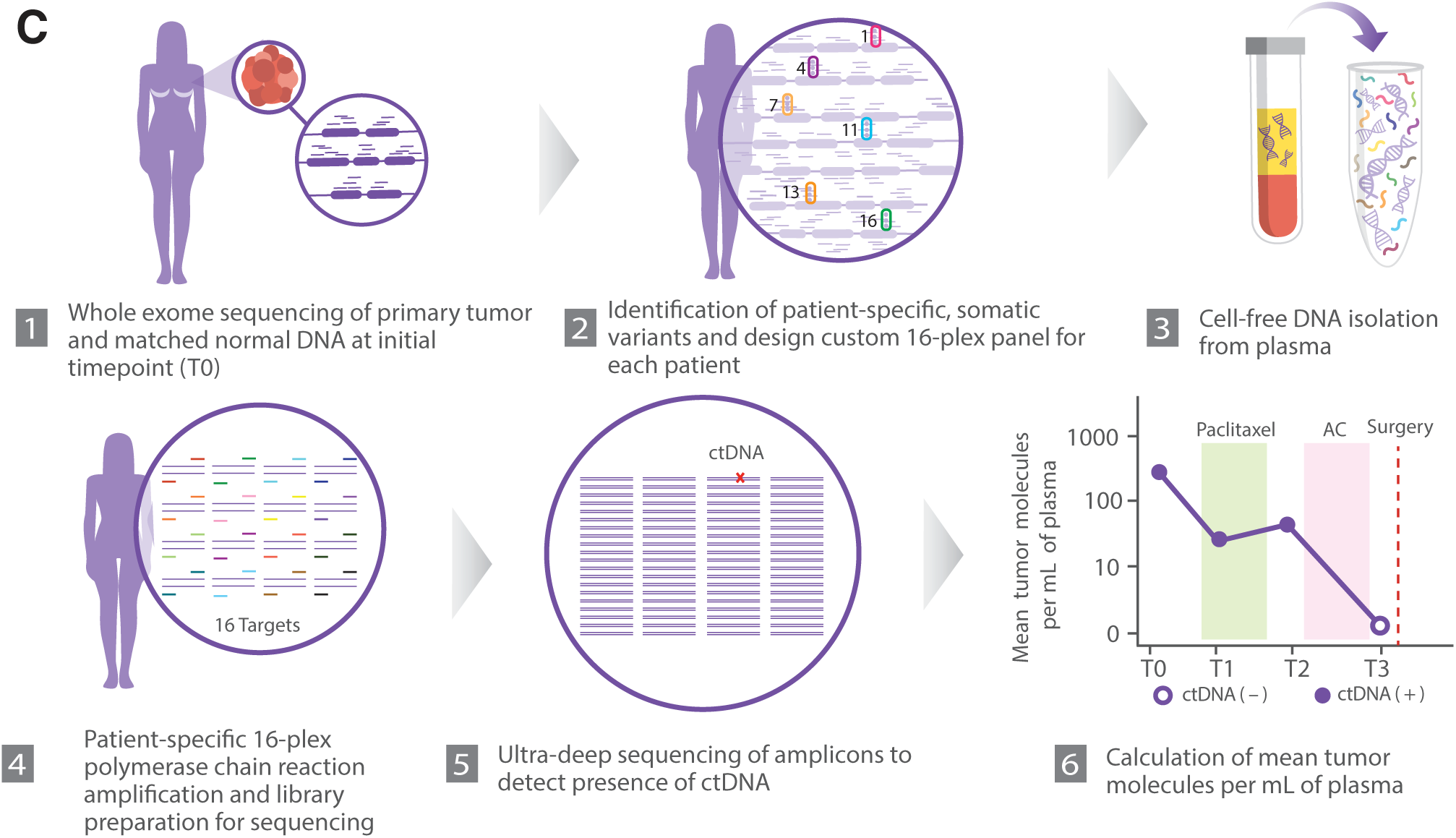
Study schema, methods for ctDNA analysis, patients and samples. **A)** Diagram showing the study schema of the I-SPY 2 TRIAL. Prior to study entry, tumor biopsy from each patient is analyzed to assess hormone-receptor (HR) and human epidermal growth factor receptor 2 (HER2) status and MammaPrint scores. Blood samples are collected at the following time points: T0-baseline/pretreatment, T1-3 weeks after initiation of therapy, T2-between two treatment regimens (paclitaxel +/- MK-2206 and anthracycline (AC)), T3-after neoadjuvant chemotherapy (NAC) prior to surgery. **B)** Flow chart showing patients and samples evaluated in the study and sample performance at different quality control (QC) points. **C)** Schema of the methods for ctDNA analysis. PCR-polymerase chain reaction.

CfDNA was isolated from plasma samples collected from pretreatment (T0), 3 weeks after initiation of treatment (T1), between paclitaxel and anthracycline regimens (T2), and after NAC prior to surgery (T3) (**Fig 1A, Supplementary Fig S2**). From the list of variants derived from WES, a unique personalized panel consisting of 16 highly ranked somatic SNV were selected (**Fig 1C, Supplementary Methods**) and used to interrogate cfDNA using 16-plex polymerase chain reaction. Amplicons were subjected to ultra-deep sequencing to detect ctDNA (**Supplementary Fig S3**). ctDNA analysis was successfully performed on 291 (87%) of the potential 336 total plasma samples (84 patients × 4 time points). Samples with at least two detectable SNVs were considered ctDNA-positive *[10, 25-27]*.

Of the 84 patients, 35% were hormone receptor-positive (HR+)/human epidermal growth factor receptor 2-negative (HER2-), 23% HER2+, and 43% triple negative breast cancers (TNBC); 30% had T3 or T4 tumors; 53% were node-negative and 61% were considered to be MammaPrint High 2 (ultra-high risk) (**Supplementary Table S1**).

### Baseline ctDNA is associated with tumor burden and aggressive phenotype

At pretreatment (T0), 73% of the patients had detectable ctDNA (**Fig 2A**). ctDNA detection rates in patients who received standard NAC (n=27, 73%) was similar to those who received additional MK-2206 (n=57, 72%) (**Fig 2A, Supplementary Table S1**). The proportion of ctDNA-positive samples was significantly higher among HER2+ (84%) and TNBC (86%) subtypes as compared to HR+/HER2-(48%) subtype (p<0.01, **Supplementary Table S1, Fig 2A-B**). ctDNA positivity was also associated with larger tumors (T3/T4, 91%, P=0.014) but not with nodal status at the time of diagnosis. A significantly higher proportion of MammaPrint High 2 patients were ctDNA-positive (86%) compared to 52% in MammaPrint High 1 (p< 0.01).

**Fig 2.**
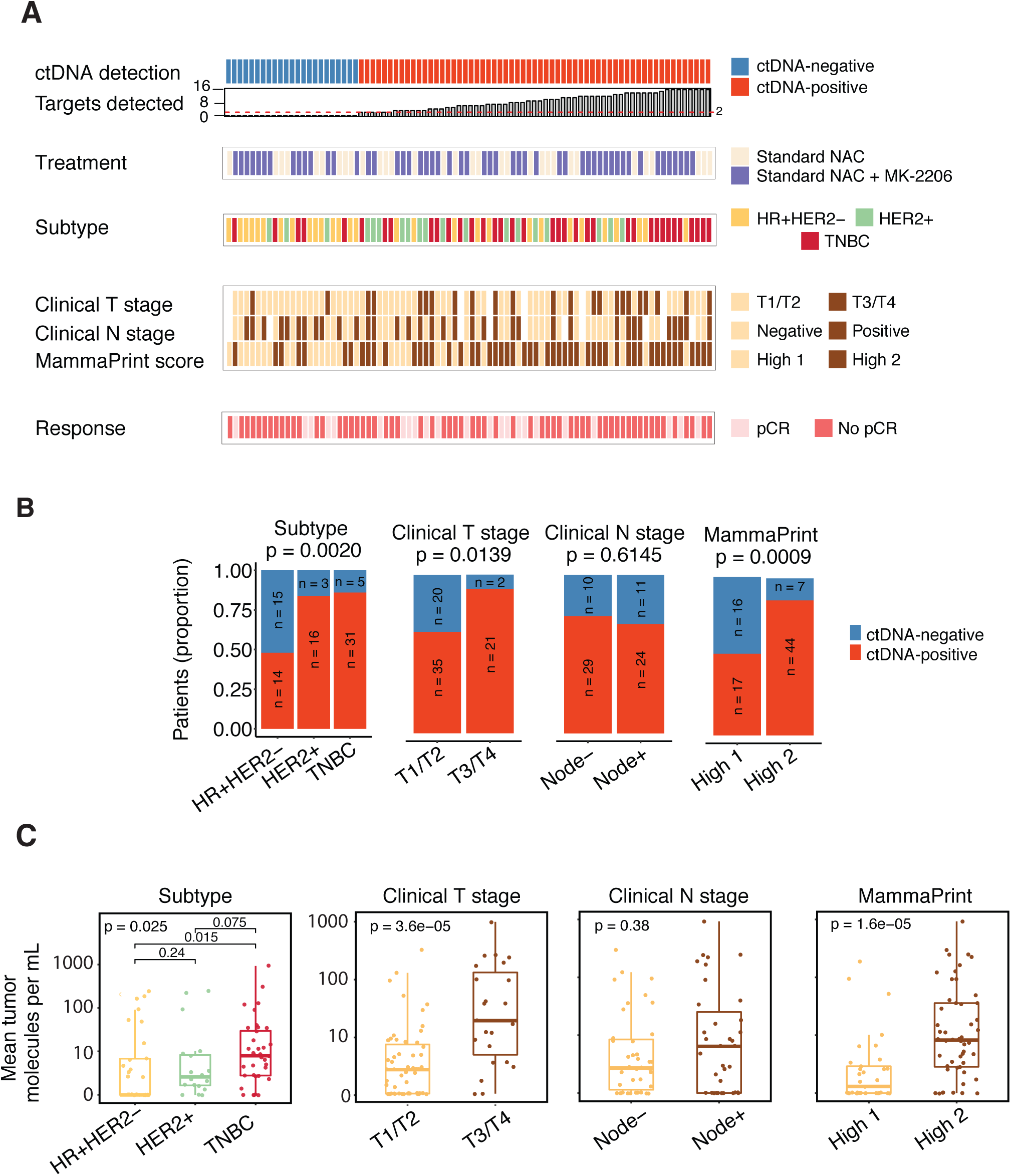
Association between ctDNA and clinicopathologic characteristics. **A)** Overview of patient and tumor characteristics according to ctDNA status at baseline (T0). HR-hormone receptor, TNBC-triple negative breast cancer, pCR-pathological complete response. **B)** Proportion of ctDNA-positive and negative patients at baseline (T0) according to clinical characteristics. P values were calculated using Fisher’s exact test. **C)** Mean tumor molecules per mL of plasma according to clinical characteristics. Distributions were compared using Wilcoxon rank sum (binary variable) or Kruskal Wallis (ternary variable) tests.

We also evaluated the absolute ctDNA levels (i.e., mean tumor molecules per mL of plasma) in the different groups stratified according to these same clinical variables and observed the same trend: Mean tumor molecules per mL in TNBC and HER2+ patients was significantly higher compared to that of HER+/HER2-patients, similarly for clinical T-stage and MammaPrint index (**Fig 2C**).

### ctDNA positivity decreases with distinct dynamics during NAC

In the population as a whole, ctDNA positivity decreased during the course of NAC, from 73% before treatment (T0), to 35% at 3 weeks (T1), to 14% at the inter-regimen time point (T2), and down to 9% after NAC (T3) (**Fig 3A**). Similarly, the absolute ctDNA levels decreased over time (**Fig 3B**). Although, on average the ctDNA positivity decreased with time, at the individual patient level, five main patterns were observed. **Fig 3C** shows ctDNA positivity as a function of time during treatment for 58 of the 84 patients who had complete serial data available at all four time points: Patients with undetectable ctDNA at T0 who remained undetectable at T3 (n=20, 34%), patients who respectively, cleared at T1 (n=20, 34%), at T2 (n=9, 16%), or at T3 (n=4, 7%), or patients who remained ctDNA-positive after NAC (T3) (n=5, 9%).

**Fig 3.**
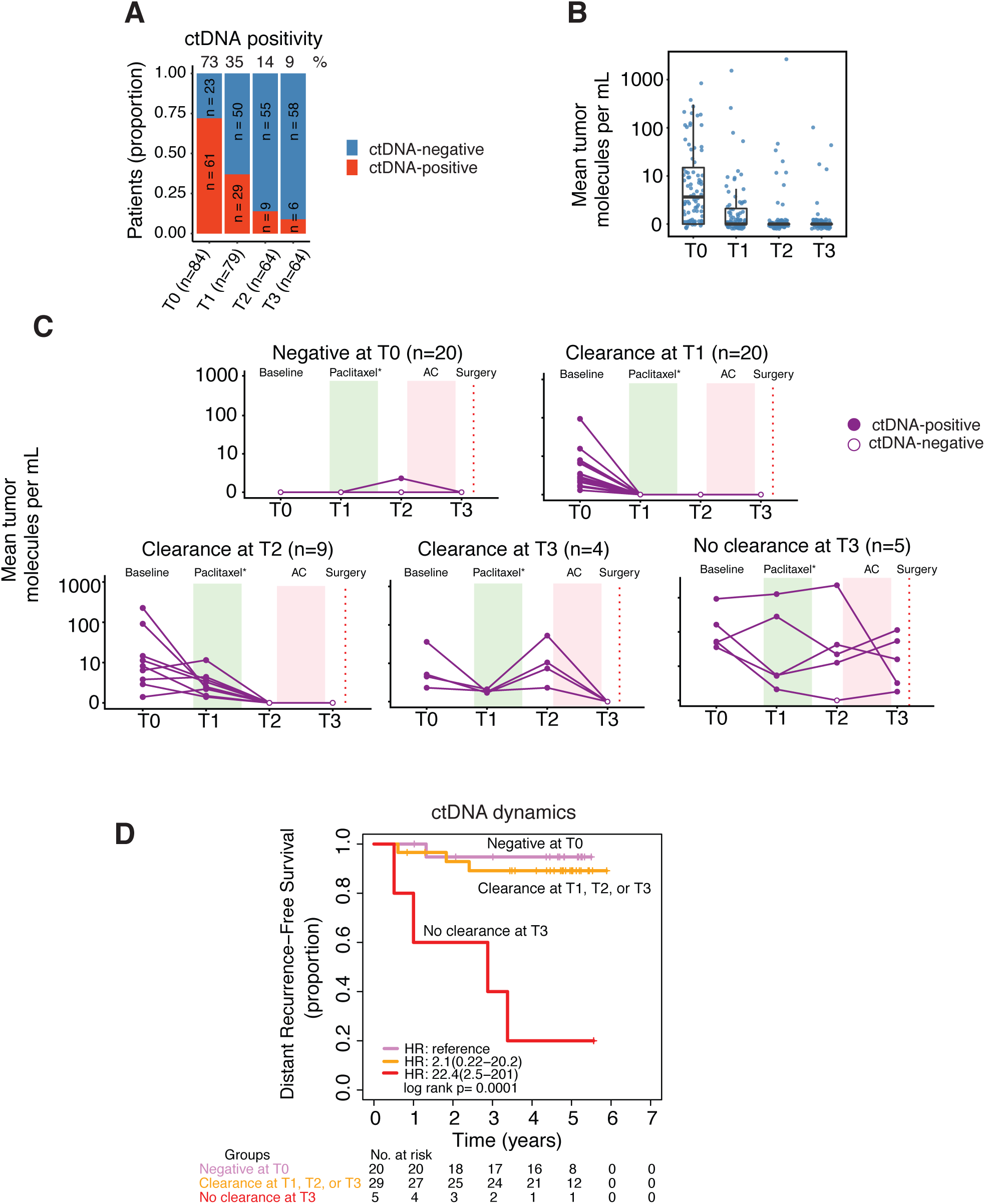
ctDNA dynamics over the course of neoadjuvant chemotherapy. **A)** Proportion of patients according to ctDNA positivity based on number of samples available per time point. **B)** Mean tumor molecules per mL of plasma across time points. **C)** Patients with complete ctDNA data for four time points (n=58) grouped according to observed patterns of ctDNA clearance or non-clearance. **D**) Survival in patients grouped according to ctDNA clearance. Of the 58 patients, 54 had survival data. Patients who cleared ctDNA at T1, T2 or T3 were combined into one group and their survival was compared with those of patients who did not clear ctDNA at T3 and those who were ctDNA-negative at T0 (reference group).

### Clearance dynamics of ctDNA is associated with NAC response

We evaluated ctDNA clearance as a predictor of response to NAC. 56 patients who were ctDNA-positive at T0 had a corresponding T1 plasma measurement (**Fig 4A)**; and of these, 29 (52%) remained ctDNA-positive at T1, 3 weeks after the initiation of treatment. 83% of patients who did not clear their ctDNA at T1 had residual disease at surgery (24/29 non-pCR) compared to 52% in patients who cleared ctDNA at T1 (14/27 non-pCR). This association was significant (OR 4.33, P=0.012, adjusted for subtype and treatment received). The positive predictive value (PPV) of the test (for predicting non-pCR) increased with time (**Fig 4B**).

**Fig 4.**
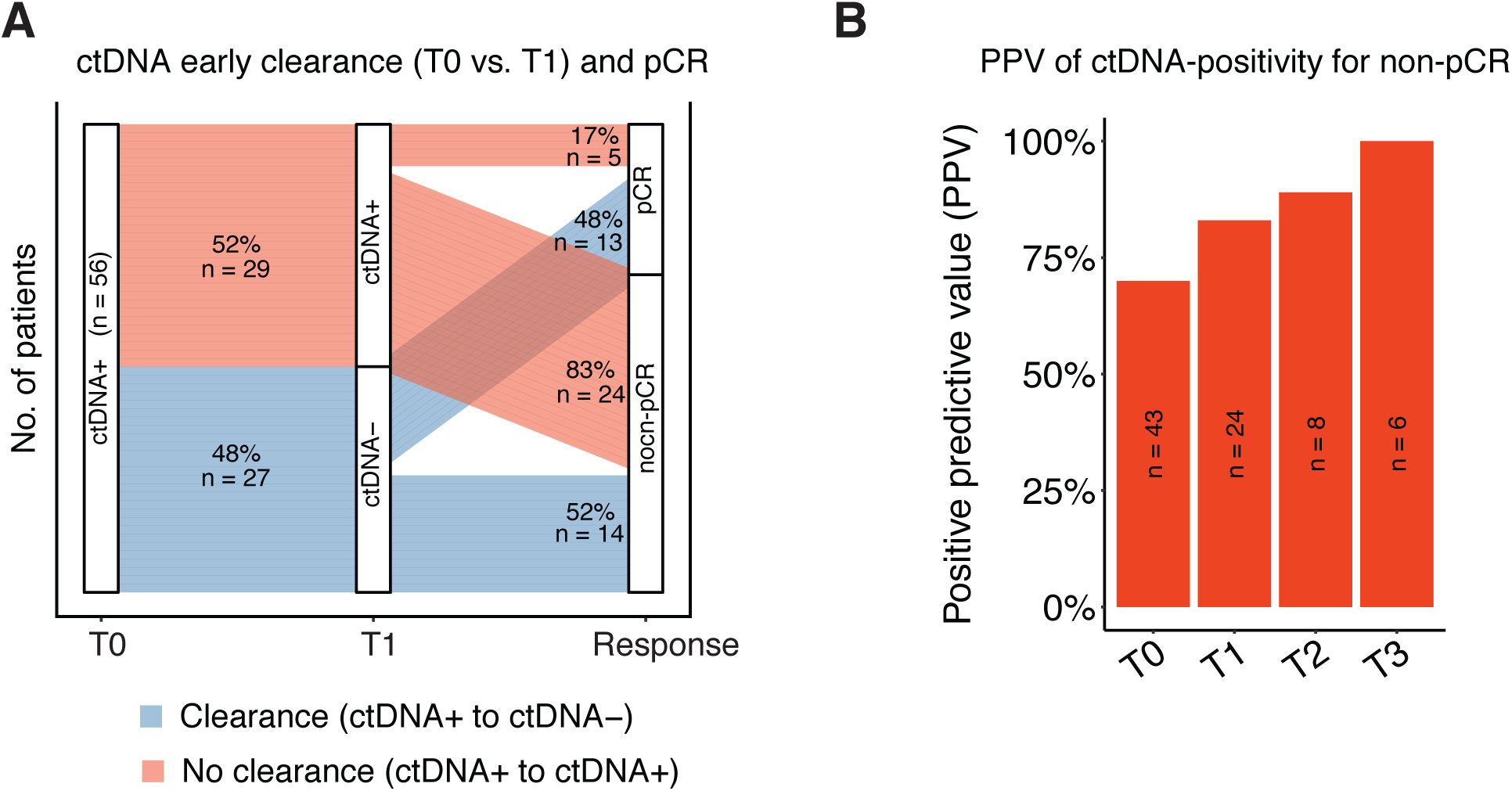
Association of ctDNA with response to neoadjuvant chemotherapy and its positive predictive value. **A)** Sankey plot showing ctDNA dynamics (clearance or non-clearance) early during treatment vs. response (pathologic complete response, pCR, or no pCR). Analysis was focused on patients who were ctDNA-positive at baseline (T0) and had corresponding ctDNA testing results at T1, 3 weeks after initiation of therapy. **B)** Positive predictive value (PPV) of ctDNA testing in predicting failure to achieve pCR (residual cancer after NAC).

### Clinical events are frequent in patients with detectable ctDNA

Survival data was available for 75 of the 84 patients, with a median follow-up of 4.8 years (range: 0.5 to 6.3 years). In this period, 8 had local recurrences, 10 experienced distant metastases, of whom 8 died (**Fig 5A**). Detectable ctDNA in at least one time point was observed in 6 of the 8 patients (75%) with local recurrence, 9 of the 10 patients (90%) who had distant recurrence and in all 8 patients who died (100%). Of note, one patient who experienced a distant recurrence but did not have detectable ctDNA was diagnosed with brain only metastasis. The lack of ctDNA detection in this type of metastasis is consistent with findings from previous studies [7, 29].

**Fig 5.**
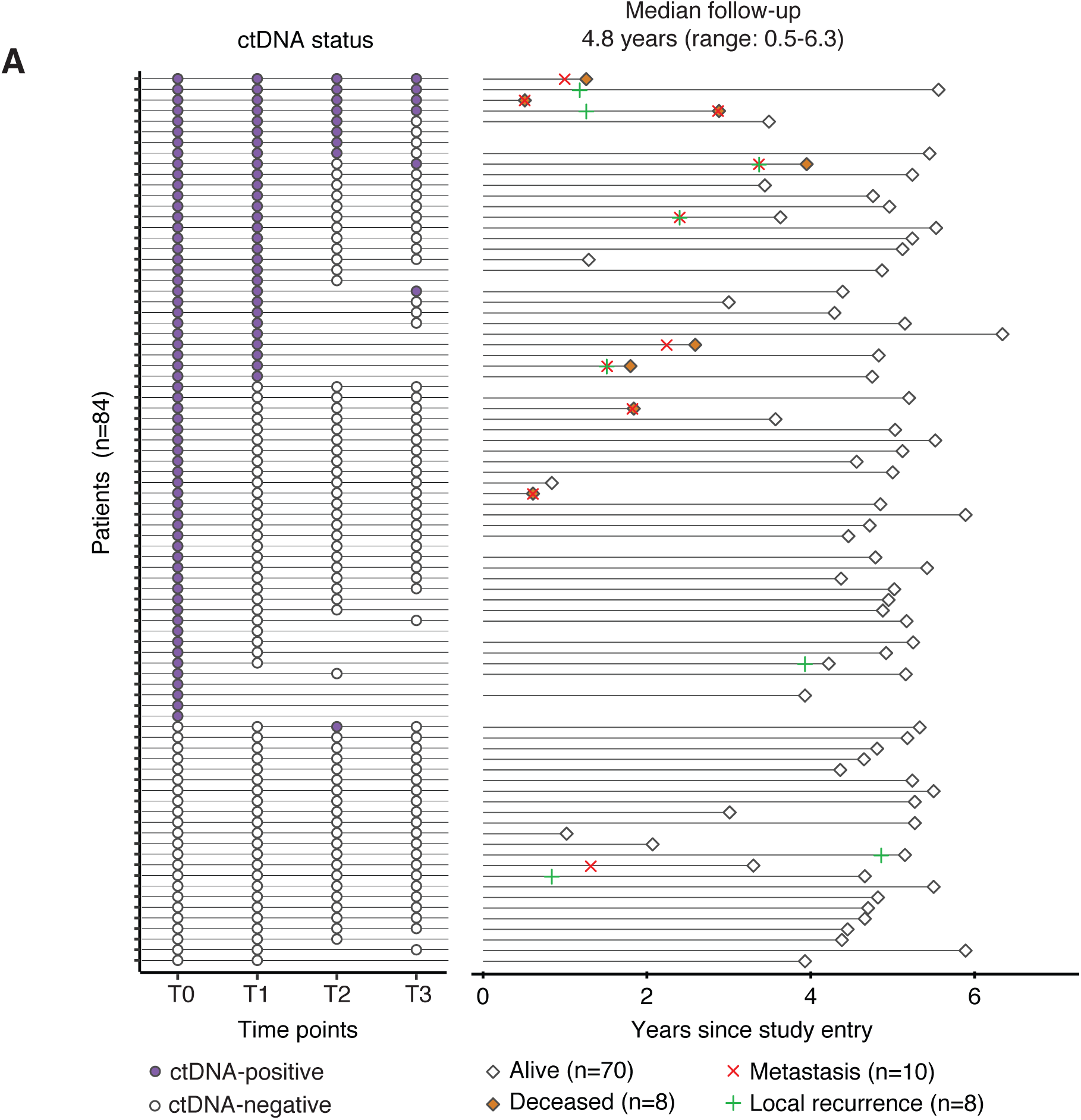
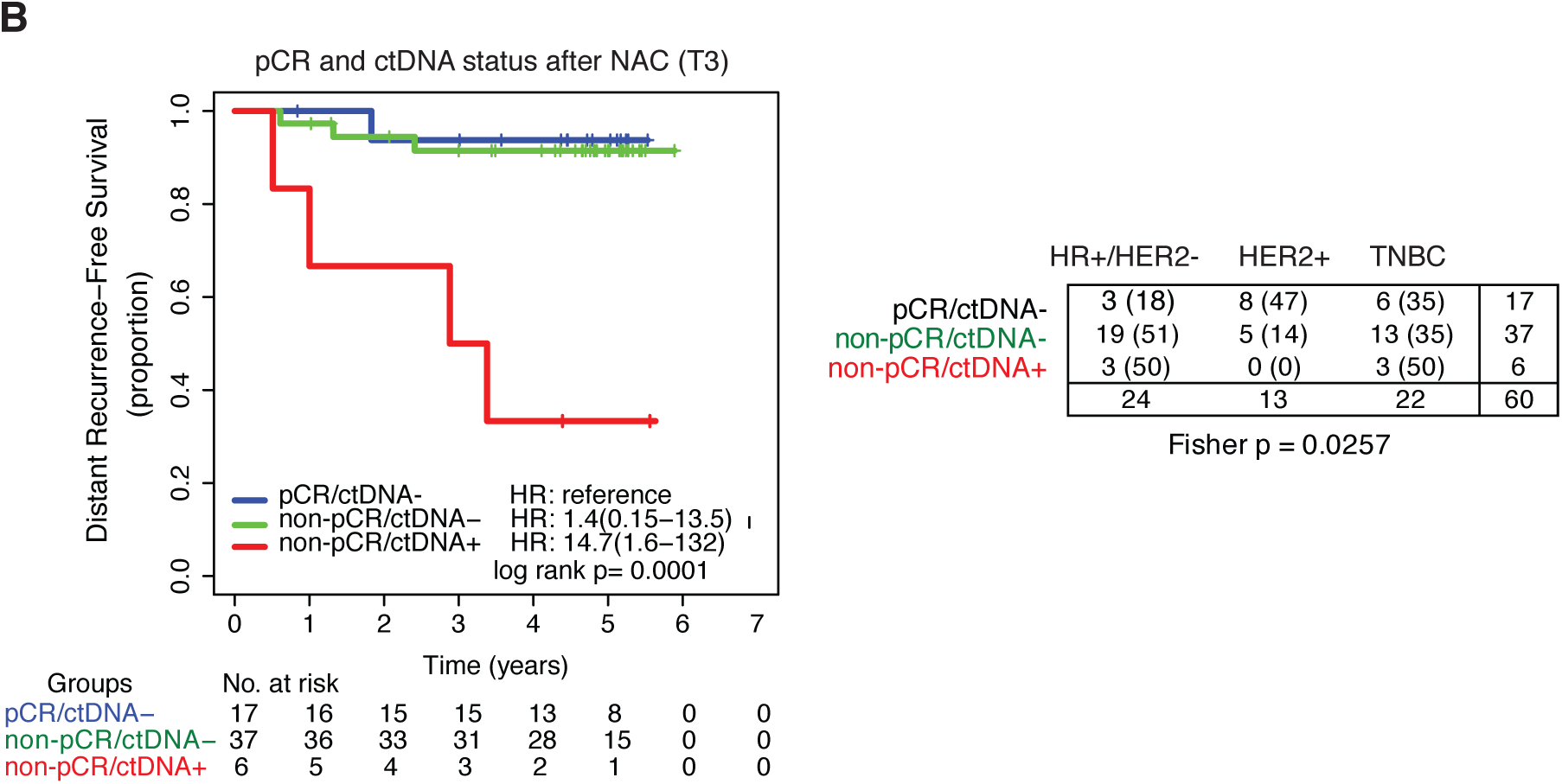
ctDNA and clinical outcomes. **A)** Overview of the ctDNA detection across different time points [T0: baseline/pretreatment, T1: 3 weeks after initiation of therapy, T2: between two treatment regimens (paclitaxel and AC), T3: after NAC prior to surgery]. The right panel shows a swimmer plot depicting the length of follow-up and events in 75 patients with survival data. The primary endpoint of the study was distant recurrence-free survival. **B)** Patient survival stratified based on ctDNA status after NAC (T3) and response to treatment (pathological complete response, pCR). Inset table shows the numbers and percentages of patients according to subtype and response/ctDNA status.

### ctDNA dynamics is significantly associated with metastatic recurrence

We examined whether ctDNA dynamic patterns (see **Fig 3C**) were associated with DRFS, the secondary endpoint of the I-SPY 2 TRIAL. Of 58 patients with ctDNA data at all time points, 54 had follow-up information. Patients who had cleared ctDNA at T1, T2 or T3 (n=29) had similar risk of metastatic recurrence compared to those who were ctDNA-negative at T0 (n=20) (hazard ratio (HR), 2.1; 95% confidence interval (CI), 0.22-20.2) (**Fig 3D**). Patients who did not clear ctDNA at T3 (n=5) had significantly higher risk of metastatic recurrence (HR, 22.4; 95%, CI, 2.5-201, p<0.001)

### ctDNA at T1, T2, and T3 but not T0 is associated with increased risk of metastatic recurrence

Next, we examined whether ctDNA status (positive or negative) at different time points was associated with DRFS (**Supplementary Fig S4**). At baseline (T0), ctDNA-positive patients had increased risk of metastatic recurrence, but this association did not reach statistical significance (HR, 4.11; 95% CI, 0.52-32.4). In contrast, ctDNA positivity at 3 weeks after initiation of therapy (T1; HR, 4.5; 95% CI, 1.2-17.4), between regimens (T2; HR, 5.4; 95% CI, 1.3-22.5), and after NAC (T3; HR, 11.5; 95% CI, 2.9-46.1) was significantly associated with increased risk of metastatic recurrence.

### Clearance of ctDNA after NAC (T3) regardless of pCR status is associated with improved survival

Patients were stratified according to their ctDNA status at T3 (after NAC) and pCR status (n=60). All 17 patients who achieved a pCR (100%) were ctDNA-negative (pCR/ctDNA-). These patients, as expected, had favorable DRFS (**Fig 5B)**. Of the 43 patients who did not achieve pCR, 86% were ctDNA negative (non-pCR/ctDNA-, n=37) and 14% were ctDNA-positive (non-pCR/ctDNA+, n=6). Of the six patients who were positive at T3, four out of six patients received MK-2206 in addition to standard NAC; three were HR+HER2-, and three had TNBC (see also **Supplementary Fig S5)**. Patients who were non-pCR/ctDNA+ had a significantly increased risk of metastatic recurrence compared to non-pCR/ctDNA-patients (HR 10.4, 95% CI, 2.3-46.6). Interestingly, the risk of metastatic recurrence in non-pCR/ctDNA-patients was similar to those who achieved a pCR (HR 1.4, 95% CI, 0.15-13.5).

## Discussion

In this study, we examined the role of personalized ctDNA as a predictive biomarker for response and outcome in the neoadjuvant setting. The cohort included early stage breast cancer patients with high-risk of recurrence and who were treated in the I-SPY 2 TRIAL.

ctDNA studies in neoadjuvant setting in breast cancer have recently been reported [28, 30-32]. Two of the four studies were limited to a particular subtype (i.e., only TNBC [28] or only HER2+ [30]). Rothé and colleagues observed that ctDNA detection before NAC was associated with decreased likelihood of achieving a pCR [30]. McDonald and colleagues showed that non-responding patients have higher ctDNA levels after NAC compared to those achieved a pCR [31]. Two of the studies examined association between ctDNA and survival [28, 30], but none were able to demonstrate the prognostic impact of residual ctDNA after NAC.

We report on the use of a personalized ctDNA test informed by each patient’s tumor genotype. We found that ctDNA is frequently detected in untreated high-risk early stage population (∼70% of patients). The patterns of change in ctDNA during NAC were significantly correlated with risk of metastatic recurrence. We also found that ctDNA testing early during NAC (at 3 weeks) provided actionable information: Persistent ctDNA identified patients who were unlikely to achieve a pCR, whereas clearance predicted improved response. These findings, in conjunction with changes in imaging may facilitate decisions on whether to switch to a more effective therapy or continue with treatment.

Our study showed for the first time that ctDNA status after NAC can further stratify patients in the non-pCR group and predict metastatic recurrence of breast cancer (**Fig 5B**). Residual ctDNA significantly correlated with increased risk of early metastatic recurrence. Most importantly, we found that clearance of ctDNA after NAC was associated with improved survival regardless of pCR status. If validated in a larger cohort, these findings could potentially change management in the neoadjuvant setting. We are currently profiling additional patients in the I-SPY 2 TRIAL, treated with other investigational drugs, including neoadjuvant immunotherapy (e.g., pembroluzimab) [33].

A number of technologies for detection of ctDNA have been developed and are described in detail in a recently published review [34]. Our approach provides several advantages over other methods of ctDNA analysis. The upfront WES of primary tumors enables personalized selection of ctDNA targets that is independent of driver status. In this study, driver mutations were not detected in 12 (14%) of the pretreatment primary tumors, and therefore, would have not been amenable to ctDNA monitoring using a pre-determined gene panel composed of driver genes. Our assay simultaneously tracks 16 patient-specific somatic variants and thus offers a more robust representation of the heterogeneity of a patient’s tumor [10, 26, 27]. In contrast, other methods like droplet digital PCR (ddPCR) [8] or BEAMing [35] track only one to a few somatic variants. Most importantly, the very deep coverage (∼95,000X depth of read for each target) allows for a low limit of detection at <0.01% VAF, [25]. Our ctDNA test does have certain limitations including the inability to detect new second primary cancers which are often genetically unrelated to the original cancer [36]; also, it will miss novel somatic variants that arise during tumor evolution in response to therapy-mediated selection pressures [37].

In the light of our findings, novel paradigms for ctDNA-directed treatment can be envisioned in future clinical trials. The current I-SPY 2 schema provides patients a single therapeutic opportunity to achieve a pCR [38] (**Fig 1A**). In the next iteration of the trial, patients will be given options to receive additional treatment to improve their chances of achieving a pCR, i.e., if the initial agent does not result in a predicted complete response. Our data suggest that ctDNA analysis can play a role in improving the identification of patients who are candidates for escalation or de-escalation of treatment. For example, the decision to switch therapy for patient without an early clinical or imaging response to a novel therapeutic agent would be supported if the patient fails to clear ctDNA. On the other hand, patients who clear their ctDNA could continue treatment. If clearance of ctDNA is confirmed as a predictor of low risk of metastatic recurrence, such information will support treatment de-escalation.

In summary, our study shows promise that an accurate and early response prediction by highly sensitive ctDNA analysis may facilitate a timely and judicious change in treatment to improve patients’ chances of achieving favorable long-term outcomes. The I-SPY 2 TRIAL provides an excellent platform to investigate how personalized ctDNA testing can complement imaging [39] and pathologic evaluation [40] of tumor response to fine-tune pCR as a surrogate endpoint for improved survival. Response over time as measured by imaging and ctDNA in the setting of early (pCR) and late (DRFS) outcomes will provide a robust framework for elucidating the potential clinical utility of ctDNA in the neoadjuvant setting.

## Materials and Methods

### Patients and study schema

This correlative study involved women with high-risk stage II and III early breast cancer who were enrolled in the multicenter neoadjuvant I-SPY 2 TRIAL (NCT01042379). Detailed descriptions of the design, eligibility, and study assessments in the I-SPY 2 TRIAL have been reported previously [22, 41]. Institutional Review Boards at all participating institutions approved the protocol. All patients signed informed consent to allow research on their biospecimen samples.

### ctDNA analysis

#### Custom panel design

Patient-specific somatic variants were identified by WES of the primary tumor and matched germline DNA samples (**Supplementary Fig S1**, and for further details see **Supplementary Information**). Observed VAF in tissue and sequence context of each genomic aberration were used to prioritize somatic SNVs and short insertion-deletions for each tumor. For each patient, 16 highly ranked variants were selected for the custom patient-specific panel. An amplicon design pipeline was used to generate multiplex PCR primer pairs for each patient-specific set of variants [10, 25-27].

#### Plasma multiplex-PCR and sequencing workflow

cfDNA extraction, quantification, library preparation, provenance testing (**Supplementary Fig S2**), next generation sequencing (NGS) workflow, and bioinformatics pipeline are described in the **Supplementary Methods**. In brief, an aliquot of each cfDNA library was used as input into the associated patient-specific 16-plex PCR reaction. The resulting 16-plex amplicon product was uniquely barcoded, followed by pooling and ultra-high depth NGS. Sequencing was performed on an Illumina HiSeq 2500 Rapid Run with 50 cycles of paired-end reads using the Illumina Paired End v2 kit with an average read depth of >90,000x per amplicon (**Supplementary Fig S3**). A previously validated cutoff of *≥*2 variants detected was used as criteria for ctDNA positivity [10, 25-27]. The cutoff was chosen based on a previously defined confidence threshold necessary to achieve high specificity of >99.8% while maintaining high sensitivity [10].

### Statistical analysis

Logistic regression was used to assess association between pCR and ctDNA clearance. Survival curves were generated by Kaplan-Meier analysis and compared using log rank test. Cox regression analysis was used to estimate HR and 95% CI. Survival data was available for 75 of the 84 patients with a median follow-up time of 4.8 years. Detailed description of the statistical methods can be found in the **Supplementary Methods**.

## Data Availability

Data will be available upon request from the I-SPY 2 TRIAL Consortium.

## Acknowledgments

The authors thank the patients and their families and the I-SPY 2 Trial Investigators for their participation in this study.

## Author Contributions

- Conception and design: C.H. J. Lin, L.J. Esserman, L.J.V. Veer
- Development of methodology: R. Salari, H. Sethi, H.T. Wu, H. Pawar, B.G. Zimmermann
- Acquisition of data: L. Brown-Swigart, G.L. Hirst, A.J.Chien, A. DeMichele, M.C. Liu, S. Asare, H. Sethi, A.S. Tin, S.V. Shchegrova, L.J. Esserman
- Development of statistical plan: M.J.M. Magbanua, C. Yau, D.M. Wolf, C.H. J. Lin
- Analysis and interpretation of data: M.J.M. Magbanua, C. Yau, D.M. Wolf, A.L. Delson, R. Salari, H. Sethi, H.T. Wu, A.S. Tin, S.V. Shchegrova, A. Aleshin, M.C. Louie, L.J. Esserman, and L.J.V. Veer
- Writing, review, and/or revision of the manuscript: M.J.M. Magbanua, L. Brown-Swigart, C. Yau, D.M. Wolf, M.C. Louie, H.T. Wu, R. Salari, A. Aleshin, C.H.J. Lin, B.G. Zimmermann, L.J. Esserman, and L.J.V. Veer
- Study supervision: M.C. Louie, A. Aleshin, C.H.J. Lin, B.G. Zimmermann, P. Billings, L.J.V. Veer
- Led the translational aspects of the study: M.J.M. Magbanua, P. Billings, A. Aleshin, M.C. Louie, L.J.V. Veer
- Advocacy involvement: A.L. Delson

